# Prevalence and associated factors with mental health outcomes among interns and residents physicians during COVID-19 epidemic in Panama: a cross-sectional study

**DOI:** 10.1101/2021.03.26.21254435

**Authors:** Edward A. Espinosa-Guerra, Edgar R. Rodríguez-Barría, Christl A. Donnelly, Jean-Paul Carrera

**Affiliations:** Public Health and Preventive Medicine, Residency Program, Complejo Hospitalario Dr. Arnulfo Arias Madrid, Panama City, Panama; School of Public Health, University of Panama, Panama City, Panama; Department of Psychiatry, Residency Program, Complejo Hospitalario Dr. Arnulfo Arias Madrid, Panama City, Panama; Department of Infectious Disease Epidemiology, MRC, Centre for Global Infectious Disease Analysis (MRC-GIDA), Imperial College London, London, United Kingdom; Department of Statistics, University of Oxford, Oxford, United Kingdom; Department of Zoology, University of Oxford, Oxford, United Kingdom; Department of Virology and Biotechnology, Gorgas Memorial Institute of Health Studies, Panama City, Panama

## Abstract

**Background:** A new coronavirus SARS-CoV-2 was associated with a newly identified respiratory syndrome, COVID-19 in Wuhan, China, in early December 2019. SARS-CoV-2 rapidly spread across the globe resulting in 117 million cases and 2.59 million deaths by March 2021. Rapidly increased numbers of COVID-19 cases overwhelmed public health systems across the world, imposing increased working hours and workloads for health care workers. Here, we have evaluated the prevalence of health outcomes and associated factors of interns and resident physicians in Panama.

**Methods:** A cross-sectional study was undertaken during July 23, 2020, to August 13, 2020, to evaluate the prevalence of health outcomes and associated factors in interns and residents across Panama. Snowball sampling was used to recruit participants. Then an electronic questionnaire with scales to evaluate anxiety disorders (GAD-7), depression (PHQ-9) and post-traumatic stress (IES-R) was evaluated. In addition, socio-demographic variables, clinical history of mental disorders and COVID-19 exposure were evaluated. Independent analyses for each mental health outcome were undertaken using a logistic regression analysis.

**Results:** A total of 517/1205 (42.9%) interns and residents were nationwide recruited. Of these 274 (53.0%) were interns and 243 (47.0%) residents. The overall prevalence of depression symptoms was 25.3%, 13.7% anxiety and 12.2% post-traumatic stress. At least, 9.3% participants reported having suicidal ideation.

The most parsimonious model showed females had a higher prevalence of mental health disorders, in all results and the married participants were more likely to present depression (OR, 1.73; 95% CI, 1.03-2.91; P = 0.039) or at least one alteration to mental health (OR, 1.66; 95% CI, 1.03-2.68; P = 0.039). Resident physicians in surgical specialties were less likely to have post-traumatic stress (OR, 0.20; 95% CI, 0.06-0.63; P = 0.006) or at least one mental health disturbance (OR, 0.46; 95% CI, 0.26-0.83; P = 0.010). A history of psychological trauma and psychiatric pathology were risk factors for most of the disorders investigated.

**Conclusions:** A high prevalence of mental health disorders was found, showing the need to mitigate this emotional burden among healthcare workers in the current context of the COVID-19 pandemic.

## Background

The global health emergency caused by the coronavirus disease (COVID-19) that emerged in the city of Wuhan, capital of Hubei province in China, in late 2019 has posed one of the greatest challenges to frontline health care workers ^1^. In Panama, COVID19 was detected on early March, 2020, although the country rapidly implemented control and mitigation strategies, Panama has one of the highest cumulative case incidences in the Americas ^2^. By July 7^th^, 2020, COVID-19 seroprevalence in Panama was estimated around 11.7% in health care worker^3^.

As countries have adopted draconian social distancing measures to reduce infections, there has been an increase in stressors that will likely increase the risk of psychiatric illness associated with COVID-19 controls in the coming months ^4^. In health care workers, the workload, the scarcity of personal protective equipment and the fear of contagion to family and friends are added to the risk for those who are in the front line of patient care, potentially triggering anxiety, fatigue, depression, irritability, fear, insomnia, etc. However, despite the efforts to comply with the provisions of governments to control the epidemic; it has been seen in studies related to severe acute respiratory syndrome (SARS) in 2003, that although affected people and health workers were motivated to comply with quarantine to reduce the risk of infecting others, emotional distress tempted some to consider violating disease control measures ^5^.

It is evident that the pandemic has led to an increase in the prevalence of mental health disorders in the world’s population ^6^. Health care workers do not escape from this reality, so it is necessary to know the factors that influence these findings, evaluating the association or not of this outcome to mental health in order to make a comprehensive approach to support those who are in the first line of care.

Intern physicians carry out visits, prepare clinical histories, evaluate the daily evolutions of the clinical condition of patients, attend teaching meetings, among other responsibilities, under the supervision of a medical officer. While the responsibilities of resident physicians are established according to each medical residency program in their training hospital, but in general, they admit patients, undertake rotations in different medical specialties, conduct daily patient evaluations, among other responsibilities.

In Panama, the internship and medical residency are carried out in public hospitals nationwide and affiliated with the Federación Nacional de Médicos Residentes e Internos (FENAMERI). Being in the front line of care, these physicians spend long working hours in hospitals and care centers, performing shifts on both weekdays and weekends. This poses a greater risk of infection due to the high workloads as the public health system reaches maximum capacity and added to the shortage of personal protective equipment, potentially being a trigger for poor mental health.

## Ethical approval

The National Research Bioethics Committee of Panama approved the study protocol by resolution EC-CNBI-2020-06-74. Participants provided online informed consent prior to completing the questionnaire.

## Methods

This cross-sectional study was conducted during the COVID-19 pandemic based on a survey of resident physicians and interns working in public health facilities in Panama (potential respondents were 408 resident physicians and 797 interns) in July 2020 and affiliated with the National Federation of Resident Physicians and Interns (FENAMERI). The objective of the study was to determine the prevalence of mental health disorders and identify their associated factors related to sociodemographic characteristics, personal history, and exposure to SARS-CoV-2.

Between July 23 and August 13, a link containing an anonymous self-report questionnaire was sent, disseminated through emails and WhatsApp using a snowball technique. Approval for this study was obtained from the National Committee on Research Bioethics (CNBI) and participants’ informed consent was obtained online. The sampling period corresponded to the period of high incidence of COVID-19 infection, in accordance with high hospital occupancy at the national level. The questionnaire collected data on sociodemographic variables (sex, age, marital status, occupation), personal history (smoking, harmful use of alcohol assessed by the AUDIT-C scale ^7^, history of psychological trauma, history of psychiatric illness, family history of psychiatric illness) and exposure information (history of COVID-19 diagnosis, family member with COVID-19 diagnosis, infected colleagues, and whether he/she attended patients with COVID-19 in the last 15 days).

Mental health outcomes were symptoms of depression, generalized anxiety, and post-traumatic stress, assessed by self-report using the 9-item Patient Health Questionnaire (PHQ-9) scales ^8^, the 7-item Generalized Anxiety Disorder scale (GAD-7) ^9^, and the 22-item Impact of Events Scale (IES-R) ^10^.

Study participants were classified according to the presence or absence of these symptoms with the following cutoff points: at least 10 on the PHQ-9 questionnaire, at least 10 on the GAD-7, and at least 26 on the IES-R. Those who scored higher were considered as having a clinically significant range of mental health impairment. The cut-off points were extracted from the original articles describing each measure^8–10^ .

In order to protect the confidentiality of the participants, the data on the type of specialty are grouped into: surgical specialties (general surgery, cardiovascular surgery, ophthalmology, otorhinolaryngology, maxillofacial surgery, neurosurgery, vascular surgery, orthopedics, etc.), medical specialties (internal medicine and its subspecialties, psychiatry, physical medicine and rehabilitation, pediatrics and its subspecialties, occupational medicine, anesthesiology, preventive medicine, etc.), and diagnostic support specialties (pathology, radiology, etc.).

For the analysis of the results, frequencies and percentages were calculated for sociodemographic data, personal and family history, exposure to COVID-19 and prevalence of each mental health finding; then univariable and multivariable logistic regression models were performed to explore the association of these results.

## Statistical analysis

Independents analyses were undertaken for each mental health impairments: depression (PHQ-9 ≥ 10), generalized anxiety (GAD-7 ≥ 10), post-traumatic stress (IES-R ≥ 26), presence of at least one impairment and presence of all three impairments. In each case, the outcome variable was the presence/absence of mental health disturbances, as determined by the cutoff point. Associations between each outcome and the study variables were tested using chi-square test and Fisher’s exact tests; P<0.05 was considered significant. Associations between each outcome and the independent variables were estimated using generalized estimating equations for logistic regression models and expressed as odds ratios (OR). Interaction variables between all variables identified as significant in the univariate analysis were examined, and marginal effects of significant interaction terms were determined. Model diagnostics were performed to check for model specification erros, Multicollinearity was also explored. The most parsimonious model was obtained with variable selection of the log likelihood test ^11^. Univariable and multivariable ORs were calculated with 95% CIs. Analyses were performed in Stata/SE version 16.1 (StataCorp, College Station, TX).

## Results

The demographics of the participants are shown in Table 1. A total of 517 physicians completed the online questionnaire, of whom 243 (47%) were resident physicians and 274 (53%) were interns, for a response rate of 42.9% (517/1205). Of the participants, 48.5% were concentrated in the province of Panama, the capital of the country. Their mean age was 28 years (SD 3.1), with a majority of respondents being female (61.5%). Seventy-eight-point nine percent were single or separated. Of the respondents, 14.9% reported having been laboratory confirmed with COVID-19.

**Table 1.** Demographic characteristics of the study population.

| Characteristics | No. (%) |
| --- | --- |
| Total | 517 (100) |
| Gender |  |
| Male | 199 (38.5) |
| Female | 318 (61.5) |
| Age (mean ± SD) | 27.9 (3.1) |
| Marital status |  |
| Unmarried or divorced | 408 (78.9) |
| Married | 109 (21.1) |
| Provinces |  |
| Bocas del Toro | 11 (2.1) |
| Chiriquí | 41 (7.9) |
| Coclé | 13 (2.5) |
| Colón | 12 (2.3) |
| Darién | 3 (0.6) |
| Herrera | 8 (1.5) |
| Los Santos | 4 (0.8) |
| Panamá | 251 (48.5) |
| Panamá Oeste | 5 (1.0) |
| Veraguas | 6 (1.2) |
| Unanswered | 163 (31.5) |
| Main occupation |  |
| Intern physician | 274 (53) |
| Medical specialties (resident) | 131 (25.3) |
| Surgical specialties (resident) | 94 (18.2) |
| Diagnostic support specialties (resident) | 18 (3.5) |
| Background information |  |
| Smoking |  |
| Yes | 46 (8.9) |
| No | 471 (91.1) |
| Alcohol misuse |  |
| Yes | 209 (40.4) |
| No | 308 (59.6) |
| Psychological trauma |  |
| Yes | 67 (13) |
| No | 450 (87) |
| Psychiatric pathology |  |
| Yes | 88 (17) |
| No | 429 (83) |
| Psychiatric pathology in family members |  |
| Yes | 189 (36.6) |
| No | 328 (63.4) |
| Exposure to COVID-19 |  |
| Been diagnosed with COVID-19 |  |
| Yes | 77 (14.9) |
| No | 440 (85.1) |
| Diagnosis of COVID-19 in family members |  |
| Yes | 138 (26.7) |
| No | 379 (73.3) |
| Diagnosis of COVID-19 in colleague |  |
| Yes | 476 (92.1) |
| No | 41 (7.9) |
| Attending COVID-19 patients |  |
| Yes | 371 (71.8) |
| No | 146 (28.2) |
| Mental health disturbances |  |
| Depression<sup>1</sup> | 131 (25.3) |
| Generalized anxiety<sup>2</sup> | 71 (13.7) |
| Posttraumatic stress<sup>3</sup> | 63 (12.2) |
| Suicidal ideation<sup>4</sup> | 48 (9.3) |

\*SD Standard Deviation
<sup>1</sup>At least 10 points on the PHQ-9 questionnaire
<sup>2</sup>At least 10 points on the GAD-7 questionnaire
<sup>3</sup>At least 33 points on the IES-R questionnaire

## Prevalence of mental health disturbances

The prevalence rate of mental health disorders was: 25.3% (131/517) for depressive symptoms, 13.7% (71/517) for generalized anxiety, and 12.2% (63/517) for post-traumatic stress disorder. Of the respondents, 32.7% (169/517) presented at least one disturbance and 5.8% (30/517) presented all three mental health disturbances.

In response to item 9 of the PHQ-9 questionnaire “How often do you think you would be better off dead or would hurt yourself in some way?” 9.3% (48/517) of respondents reported having suicidal thoughts; 0.8% (4/517) reported having suicidal ideation “more than half the days” and 8.5% (44/517) reported experiencing it “several days.”

## Logistic regression models

### Univariable analysis

Female sex and personal history of psychological trauma and psychiatric pathology were associated with all three mental health outcomes investigated (Table 3). For example, depression among women: OR=1.75; 95% CI, 1.14-2.69; P = 0.010; generalized anxiety OR=3.19; 95% CI, 1.70-5.99; P <0.001; post-traumatic stress OR=2.98; 95% CI, 1.54-5.74; P = 0.001; with at least one mental health disturbance OR=1.93; 95% CI, 1.30-2.87; P = 0.001; and with all three mental health disturbances OR=4.34; 95% CI, 1.49-12.63; P = 0.007.

**Table 2.** Independent factors associated with mental health outcomes in univariate generalized estimating equations for logistic regression models (n = 517)

| Factor | Depression<sup>1</sup> |  |  | Generalized anxiety<sup>2</sup> |  |  | Post-traumatic stress<sup>3</sup> |  |  | At least one outcome |  |  | Presence of all three outcomes |  |  |
| --- | --- | --- | --- | --- | --- | --- | --- | --- | --- | --- | --- | --- | --- | --- | --- |
|  | OR | CI 95% | p | OR | CI 95% | p | OR | CI 95% | p | OR | CI 95% | p | OR | CI 95% | p |
| Gender |  |  |  |  |  |  |  |  |  |  |  |  |  |  |  |
| Male | Ref. | - | - | Ref. | - | - | Ref. | - | - | Ref. | - | - | Ref. | - | - |
| Female | **1.75** | **(1.14-2.69)** | **0.010** | **3.19** | **(1.70-5.99)** | **<0.001** | **2.98** | **(1.54-5.74)** | **0.001** | **1.93** | **(1.30-2.87)** | **0.001** | **4.34** | **(1.49-12.63)** | **0.007** |
| Age | 0.99 | (0.92-1.05) | 0.703 | 1.00 | (0.93-1.09) | 0.909 | 0.91 | (0.82-1.00) | 0.059 | 0.95 | (0.89-1.01) | 0.126 | 0.99 | (0.89-1.12) | 0.988 |
| Marital status |  |  |  |  |  |  |  |  |  |  |  |  |  |  |  |
| Unmarried or divorce | Ref. | - | - | Ref. | - | - | Ref. | - | - | Ref. | - | - | Ref. | - | - |
| Married | 1.37 | (0.86-2.19) | 0.183 | 1.00 | (0.54-1.85) | 0.992 | 0.77 | (0.39-1.53) | 0.453 | 1.32 | (0.85-2.05) | 0.218 | 0.93 | (0.37-2.34) | 0.881 |
| Main occupation |  |  |  |  |  |  |  |  |  |  |  |  |  |  |  |
| Intern physician | Ref. | - | - | Ref. | - | - | Ref. | - | - | Ref. | - | - | Ref. | - | - |
| Medical specialties (resident) | 0.73 | (0.45-1.19) | 0.209 | 0.59 | (0.31-1.12) | 0.109 | 0.56 | (0.29-1.08) | 0.088 | 0.65 | (0.41-1.01) | 0.058 | 0.76 | (0.31-1.85) | 0.542 |
| Surgical specialties (resident) | 0.62 | (0.35-1.10) | 0.105 | 0.52 | (0.25-1.12) | 0.095 | **0.23** | **(0.08-0.65)** | **0.006** | **0.45** | **(0.26-0.78)** | **0.005** | 0.44 | (0.13-1.53) | 0.198 |
| Diagnostic support specialties (resident) | 0.49 | (0.13-1.75) | 0.275 | 0.62 | (0.14-2.79) | 0.533 | 0.30 | (0.04-2.31) | 0.247 | 0.45 | (0.14-1.41) | 0.172 | 0.79 | (0.10-6.26) | 0.823 |
| Background information |  |  |  |  |  |  |  |  |  |  |  |  |  |  |  |
| Smoking | 1.65 | (0.87-3.14) | 0.126 | 1.87 | (0.88-3.95) | 0.103 | **3.77** | **(1.88-7.54)** | **<0.001** | **2.02** | **(1.10-3.72)** | **0.024** | **2.79** | **(1.08-7.23)** | **0.034** |
| Alcohol misuse | 1.46 | (0.98-2.18) | 0.063 | 0.95 | (0.57-1.59) | 0.855 | 1.21 | (0.71-2.05) | 0.488 | 1.10 | (0.76-1.60) | 0.609 | 1.51 | (0.72-3.16) | 0.274 |
| Psychological trauma | **2.42** | **(1.42-4.12)** | **0.001** | **4.00** | **(2.21-7.22)** | **<0.001** | **2.69** | **(1.42-5.09)** | **0.002** | **2.40** | **(1.43-4.04)** | **0.001** | **5.23** | **(2.39-11.45)** | **<0.001** |
| Psychiatric pathology | **2.43** | **(1.50-3.94)** | **<0.001** | **5.29** | **(3.07-9.12)** | **<0.001** | **2.64** | **(1.46-4.76)** | **0.001** | **2.88** | **(1.80-4.61)** | **<0.001** | **4.20** | **(1.96-9.01)** | **<0.001** |
| Psychiatric pathology in family members | 1.30 | (0.87-1.96) | 0.200 | **1.72** | **(1.04-2.86)** | **0.034** | **1.95** | **(1.15-3.32)** | **0.013** | 1.31 | (0.90-1.91) | 0.160 | **2.77** | **(1.30-5.89)** | **0.008** |

**Table 3.** Independent factors associated with mental health outcomes in multivariable generalized estimating equations for logistic regression models (n = 517)

|  |  |  |  |  |  |  |  |  |  |  |  |  |  |  |  |
| --- | --- | --- | --- | --- | --- | --- | --- | --- | --- | --- | --- | --- | --- | --- | --- |
| Exposure to COVID-19 |  |  |  |  |  |  |  |  |  |  |  |  |  |  |  |
| Been diagnosed with COVID-19 | 1.74 | (1.04-2.93) | 0.035 | 1.65 | (0.88-3.11) | 0.115 | 1.97 | (1.04-3.74) | 0.037 | 1.21 | (0.73-2.01) | 0.457 | 3.13 | (1.40-6.99) | 0.005 |
| Diagnosis of COVID-19 in family members | 1.23 | (0.79-1.90) | 0.357 | 1.00 | (0.57-1.77) | 0.989 | 1.83 | (1.06-3.18) | 0.031 | 1.35 | (0.90-2.03) | 0.145 | 1.19 | (0.53-2.66) | 0.673 |
| Diagnosis of COVID-19 in colleague | 2.08 | (0.85-5.06) | 0.107 | 3.30 | (0.78-14.00) | 0.105 | 1.83 | (0.55-6.10) | 0.327 | 1.80 | (0.84-3.86) | 0.131 | * | * | * |
| Attending COVID-19 patients | 1.37 | (0.87-2.16) | 0.179 | 1.18 | (0.67-2.10) | 0.561 | 0.70 | (0.40-1.22) | 0.210 | 1.23 | (0.81-1.87) | 0.325 | 1.08 | (0.47-2.50) | 0.844 |
| How difficult have these problems made it for you to do your work? |  |  |  |  |  |  |  |  |  |  |  |  |  |  |  |
| Not difficult at all | Ref | - | - | Ref | - | - | - | - | - | - | - | - | - | - | - |
| Somewhat difficult | 3.85 | (2.44-6.08) | <0.001 | 6.60 | (3.14-13.89) | <0.001 | - | - | - | - | - | - | - | - | - |
| Very / extremely difficult | 22.28 | (8.85-56.08) | <0.001 | 53.41 | (19.37-147.24) | <0.001 | - | - | - | - | - | - | - | - | - |

\*No one has answered the question with this outcome
<sup>1</sup>At least 10 points on the PHQ-9 questionnaire
<sup>2</sup>At least 10 points on the GAD-7 questionnaire
<sup>3</sup>At least 26 points on the IES-R questionnaire

Being a resident physician in surgical specialties was significantly associated with a reduced risk of post-traumatic stress (OR, 0.23; 95% CI, 0.08-0.65; P = 0.006) and of developing at least 1 mental health disorder (OR, 0.45; 95% CI, 0.26-0.78; P = 0.005).

Smoking was significantly associated with post-traumatic stress, having at least 1 disorder, and having all three mental health disorders (Table 3). On the other hand, having a history of having been diagnosed with COVID-19 increased the risk of presenting depression, post-traumatic stress and all three mental health disorders; while having a history of having a family member diagnosed with COVID-19 was significantly associated with post-traumatic stress (Table 3).

Respondents who presented depression and generalized anxiety found these symptoms made it difficult to perform their work (Table 3 and 4).

| Factor | Depression<sup>1</sup> |  |  | Generalized anxiety<sup>2</sup> |  |  | Posttraumatic stress<sup>3</sup> |  |  | At least one outcome |  |  | Presence of all three outcomes |  |  |
| --- | --- | --- | --- | --- | --- | --- | --- | --- | --- | --- | --- | --- | --- | --- | --- |
|  | OR | CI 95% | P | OR | CI 95% | P | OR | CI 95% | p | OR | CI 95% | p | OR | CI 95% | p |
| Gender |  |  |  |  |  |  |  |  |  |  |  |  |  |  |  |
| Male | Ref. | - | - | Ref. | - | - | Ref. | - | - | Ref. | - | - | Ref. | - | - |
| Female | **1.64** | **(1.03-2.61)** | **0.035** | **2.62** | **(1.30-5.28)** | **0.007** | **3.70** | **(1.77-7.71)** | **<0.001** | **1.89** | **(1.24-2.88)** | **0.003** | **3.84** | **(1.29-11.50)** | **0.016** |
| Marital status |  |  |  |  |  |  |  |  |  |  |  |  |  |  |  |
| Unmarried or divorce | Ref. | - | - | - | - | - | - | - | - | Ref. | - | - | - | - | - |
| Married | **1.73** | **(1.03-2.91)** | **0.039** | - | - | - | - | - | - | **1.66** | **(1.03-2.68)** | **0.039** | - | - | - |
| Main occupation |  |  |  |  |  |  |  |  |  |  |  |  |  |  |  |
| Intern physician | - | - | - | - | - | - | Ref. | - | - | Ref. | - | - | - | - | - |
| Medical specialties (resident) | - | - | - | - | - | - | 0.61 | (0.30-1.22) | 0.164 | 0.62 | (0.38-1.01) | 0.055 | - | - | - |
| Surgical specialties (resident) | - | - | - | - | - | - | **0.20** | **(0.06-0.63)** | **0.006** | **0.46** | **(0.26-0.83)** | **0.010** | - | - | - |
| Diagnostic support specialties (resident) | - | - | - | - | - | - | 0.58 | (0.07-4.71) | 0.614 | 0.60 | (0.18-1.95) | 0.395 | - | - | - |
| Background information |  |  |  |  |  |  |  |  |  |  |  |  |  |  |  |
| Smoking | - | - | - | - | - | - | **6.12** | **(2.71-13.81)** | **<0.001** | **2.19** | **(1.13-4.23)** | **0.020** | - | - | - |
| Psychological trauma | **1.98** | **(1.10-3.55)** | **0.022** | **2.61** | **(1.29-5.27)** | **0.007** | **2.12** | **(1.06-4.26)** | **0.034** | - | - | - | **4.01** | **(1.75-9.16)** | **0.001** |
| Psychiatric pathology | - | - | - | **3.21** | **(1.71-6.03)** | **<0.001** | **2.03** | **(1.07-3.84)** | **0.030** | **2.46** | **(1.51-4.02)** | **<0.001** | **2.98** | **(1.33-6.80)** | **0.008** |
| Exposure to COVID-19 |  |  |  |  |  |  |  |  |  |  |  |  |  |  |  |
| Been diagnosed with COVID-19 |  |  |  |  |  |  |  |  |  |  |  |  | **2.48** | **(1.05-5.83)** | **0.037** |
| Diagnosis of COVID-19 in family members | - | - | - | - | - | - | 1.92 | (1.06-3.48) | 0.031 | - | - | - | - | - | - |
| How difficult have these problems made it for you to do your work? |  |  |  |  |  |  |  |  |  |  |  |  |  |  |  |
| Not difficult at all | Ref. | - | - | Ref. | - | - | - | - | - | - | - | - | - | - | - |
| Somewhat difficult | 3.86 | (2.42-6.16) | <0.001 | 5.48 | (2.56-11.73) | <0.001 | - | - | - | - | - | - | - | - | - |
| Very / extremely difficult | 21.86 | (8.54-55.96) | <0.001 | 30.53 | (10.52-88.55) | <0.001 | - | - | - | - | - | - | - | - | - |

least 10 points on the PHQ-9 questionnaire least 10 points on the GAD-7 questionnaire least 26 points on the IES-R questionnaire

### Multivariable analysis

The most parsimonious multivariable model revealed that female sex was positively associated with the prevalence of mental health disturbances in all the outcomes analyzed (Table 4). Married participants were more likely to have depression (OR, 1.73; 95% CI, 1.03-2.91; P = 0.039) or at least one mental health disturbance (OR, 1.66; 95% CI, 1.03-2.68; P = 0.039).

Surgical specialties were less likely to have PTSD or at least one mental health disturbance, while smoking was significantly associated with self-report of these disturbances (Table 4).

Having a previous diagnosis of COVID-19 was associated with having all three mental health disorders and having a family member with COVID-19 was significantly associated with post-traumatic stress.

## Discussion

Our findings document a high prevalence of symptoms associated with mental health disorders during the first peak of COVID-19 cases in Panama. We found prevalence rates of 25.3% for depression, 13.7% for anxiety, and 12.2% for post-traumatic stress disorder, comparatively similar to those found in health personnel in Dutch ^12^ and Italian ^13^ but below those found during this outbreak in China ^14^.

In high-incidence areas, an epidemic event of this magnitude is comparable to previous events such as SARS, where a large percentage of health care workers in high-risk situations reported symptoms related to mental health disturbances ^15^.

Female respondents were 1.74 to 4.34 times more likely to report a mental health disturbance compared to males, consistent with a systematic review by Ricci et al. where women were more likely than men to report mental health disturbances, especially for schizotypal personality disorder (STPD) ^16^. The history of psychological trauma in females as a risk factor for the development of mental health disorders may be attributed to gender differences in the rates and types of trauma exposure (e.g., natural disasters, motor vehicle accidents, rape, physical attack, combat, etc.) ^17^.

Most participants were medical interns, indicating that most of them had fewer years of work experience and are probably exposed to a higher risk of infection. This study indicated that having a history of psychological trauma and psychiatric pathology was associated with experiencing depression, generalized anxiety and post-traumatic stress disorder outcomes, in all models conducted. This is consistent with the findings of Young et al. where healthcare workers with a history of mental illness were at increased risk for significant emotional symptoms ^18^.

Being married was associated with depressive symptoms, which may be due to competing family responsibilities and/or fear of partner contagion. This pattern was also observed during the SARS epidemics in 2003 in China ^19^ and Canada ^20^. On the other hand, the occupation of resident physician in surgical specialties reduced the risk of post-traumatic stress and of developing at least 1 mental health disorders. Elective surgeries were suspended in all hospitals in the country, as a preventive measure adopted by the Ministry of Health, to limit transmission risks in hospital settings ^21^.

As previous studies indicate, smoking is an important mental health issue, and in this study, smokers were 2-5 times more at risk for post-traumatic stress disorder and 2 times more at risk for at least 1 mental health disturbance than non-smokers^22^. These findings could be due to in part to the pandemic generating factors that contribute to smoking initiation or the vulnerability of people with a significant psychological history has triggered the habit of smoking ^6^.

A troubling finding immersed in the depression data is the rate of individuals reporting positive responses to item 9 of the PHQ-9 questionnaire (“How often do you think you would be better off dead or hurt in some way?”). Of the respondents, 9.3% reported having suicidal thoughts, within them; 0.8% reported having suicidal ideation half of the days and 8.5% reported experiencing it almost every day. This result indicated elevated suicidal ideation above that found in U.S. health care workers by Young et al. ^18^ and close to that reported by the CDC, with 10.7% of persons experiencing suicidal thoughts associated with the COVID-19 pandemic ^23^.

In a study to determine whether responses to the PHQ-9 questionnaire predict subsequent suicide attempt or death by suicide, Simon et al. found that those individuals with positive responses to item 9 of the PHQ-9 were six times more likely to attempt suicide and five times more likely to die by suicide within one year than those who did not report such thoughts ^24^. Another study by Rossom et al. demonstrated that patients with any level of suicidal ideation on item 9 of the PHQ-9 were almost twice as likely to attempt suicide in the following year ^25^.

Additionally, our findings indicate that exposure to COVID-19 at work or at home through the illness of a family member with COVID-19 independently contributed to presenting with all three mental health disturbances together and post-traumatic stress, respectively. Similar results were evident during the SARS epidemic in 2003 where having had a family member or friend with SARS was a strong predictor of the level of post-traumatic stress symptoms, suggesting a special feature of the psychosocial effects of infectious disease outbreaks, which can be differentiated from other disasters ^19^.

Previous studies have shown that young physicians are a vulnerable professional group requiring professional support to prevent mental health disorders due to long working hours, and support from senior physicians may have a positive effect on some of these disorders, especially in health emergencies ^26^.

Despite a response rate of 42.9%, non-response bias may exist because the remaining resident physicians and interns might have shown less interest in participating because they were not as emotionally affected by the current pandemic. Therefore, it is difficult to estimate how representative the responses are of the population of residents and interns or how biased the estimates might be due to nonresponse. On the other hand, we assume that the reliability of the responses is high because the data collected in the survey are familiar to residents and interns and they are able to recognize the mental illnesses studied here. With the aim of assessing mental disturbances at the time of response, the questions in the questionnaire focused on symptoms during the last 15 days.

## Conclusions

With a response rate of 42.9%, one third of our sample presented at least one clinically significant mental health disturbance (including suicidal ideation), with women being the most affected in all analyses.

Our findings raise concerns about the psychological well-being of resident physicians and interns involved in care during the first wave of the COVID-19 epidemic in Panama.

It is essential for institutions to intervene with those most at risk of presenting psychiatric symptoms and offer them sources of support to mitigate the emotional impact and increase resilience in situations of occupational stress.

## Data Availability

All data is available within the manuscript. Dataset are available upon request to the corresponding authors, in this case notification the the National IRB should be provided.

## List of abbreviations

SARS-CoV-2: severe acute respiratory syndrome type 2 coronavirus
COVID-19: coronavirus disease 2019
PHQ-9: Patient Health Questionnaire
GAD-7: General Anxiety Disorder-7
IES-R: Impact of Event Scale-Revised
PTSD: Post-traumatic stress disorder
COVID-19: Coronavirus Disease 2019
FENAMERI: Federación Nacional de Médicos Residentes e Internos
CDC: Center for Disease Control and Prevention

## Statements

### Data and material availability

The data set generated and analyzed in the current study is available from the corresponding author upon reasonable request.

### Funds

This work was supported by SENACYT (grant of rapid response to COVID-19 pandemic: FID-03-2021-COVID19-08) grant to J-PC. J-PC is funded by the Clarendon Scholarship from the University of Oxford and Lincoln-Kingsgate Scholarship from Lincoln College-Oxford. “This work was jointly funded by the UK Medical Research Council (MRC) and the UK Department for International Development (DFID) under the MRC/DFID Concordat agreement and is also part of the EDCTP2 programme supported by the European Union (MR/R015600/1). CAD thanks the UK NIHR for funding of the NIHR Health Protection Research Unit in Emerging and Zoonotic Infections.” EB is member of the Sistema Nacional de Investigación (SNI-SENACYT), Panamá.

### Author’s contributions

EE conceived and designed the study. EE and EB constructed the questionnaire. EE and EB collected the data. EE and J-PC performed the statistical analysis and CD advised on statistical analysis. EE, EB and J-PC wrote the manuscript. EE, EB, CD, J-PC supervised the project and contributed to the writing of the paper. All authors reviewed the manuscript. All authors read and approved the final manuscript.

## Acknowledgements

The authors would like to thank all the residents and interns physicians in Panama for their contribution to patient care during this health crisis and their participation in this study.

---

**Supplementary Table 1.**
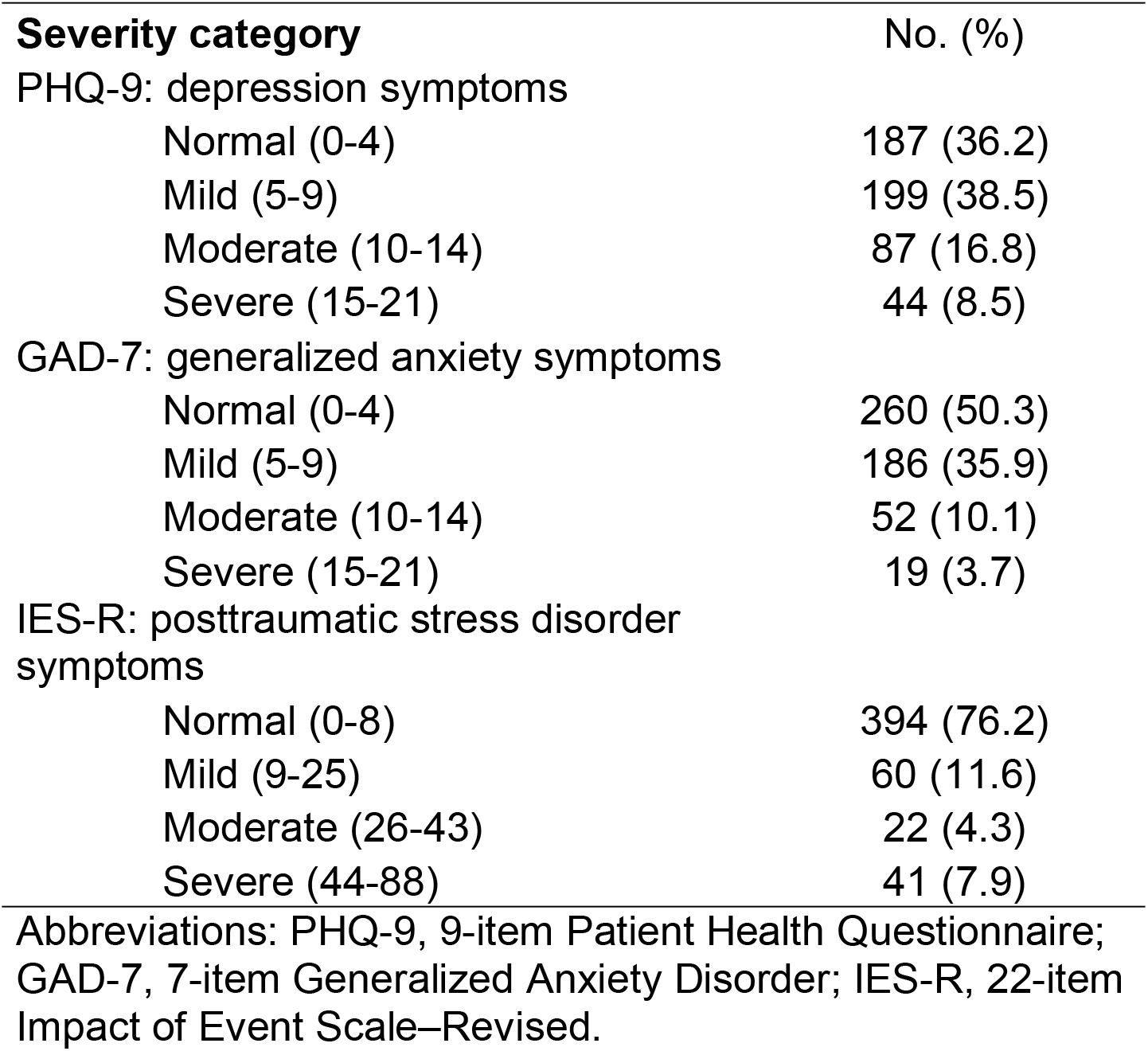
Symptom severity of depression, generalized anxiety and posttraumatic stress (n = 517)

| **Severity category** | **No. (%)** |
| --- | --- |
| PHQ-9: depression symptoms |  |
| Normal (0-4) | 187 (36.2) |
| Mild (5-9) | 199 (38.5) |
| Moderate (10-14) | 87 (16.8) |
| Severe (15-21) | 44 (8.5) |
| GAD-7: generalized anxiety symptoms |  |
| Normal (0-4) | 260 (50.3) |
| Mild (5-9) | 186 (35.9) |
| Moderate (10-14) | 52 (10.1) |
| Severe (15-21) | 19 (3.7) |
| IES-R: posttraumatic stress disorder symptoms |  |
| Normal (0-8) | 394 (76.2) |
| Mild (9-25) | 60 (11.6) |
| Moderate (26-43) | 22 (4.3) |
| Severe (44-88) | 41 (7.9) |

Abbreviations: PHQ-9, 9-item Patient Health Questionnaire; GAD-7, 7-item Generalized Anxiety Disorder; IES-R, 22-item Impact of Event Scale–Revised.

**Supplementary Table 2.** Scores of depression, generalized anxiety and posttraumatic stress in participants (n = 517)

| Scale | Total scores, mean<br>(IC 95%) | Occupation |  |  |  |
| --- | --- | --- | --- | --- | --- |
|  |  | Mean (IC 95%) |  |  |  |
|  |  | Intern physician | Medical specialties<br>(resident) | Surgical<br>specialties<br>(resident) | Diagnostic support<br>specialties<br>(resident) |
| PHQ-9: depression symptoms | 7.0 (6.5-7.4) | 7.7 (7.2-8.1) | 6.3 (5.5-7.0) | 5.9 (4.9-6.8) | 6.1 (3.6-8.7) |
| GAD-7: generalized anxiety symptoms | 5.2 (4.8-5.5) | 5.6 (5.0-6.1) | 4.8 (4.2-5.5) | 4.7 (3.8-5.6) | 4.6 (2.6-6.6) |
| IES-R: posttraumatic stress disorder<br>symptoms | 15.0 (13.8-16.2) | 17.5 (13.8-17.2) | 13.5 (11.3-15.7) | 10.1 (7.9-12.3) | 14.0 (7.8-20.1) |

Abbreviations: PHQ-9, 9-item Patient Health Questionnaire; GAD-7, 7-item Generalized Anxiety Disorder; IES-R, 22-item Impact of Event Scale–Revised.

